# Specificity of polygenic scores for psychiatric disorders beyond transdiagnostic genetic risk (p)

**DOI:** 10.1101/2025.06.12.25329497

**Authors:** Engin Keser, Wangjingyi Liao, Andrea G. Allegrini, Thalia C. Eley, Kaili Rimfeld, Margherita Malanchini, Robert Plomin

## Abstract

**Importance:** Polygenic scores are increasingly used to estimate genetic liability for psychiatric disorders. However, their limited specificity to the disorders for which they are derived limits their clinical and research utility.

**Objective:** To determine whether associations between polygenic scores for major psychiatric disorders and psychopathology outcomes are primarily driven by transdiagnostic (*p*) or disorder-specific genetic risk.

**Design:** Cross-sectional study. Analyses were conducted from May 2024 to October 2024.

**Setting:** Population-based sample from the Twins Early Development Study.

**Participants:** Twins born in England and Wales between 1994 and 1996. Mental health data were collected from 2021 to 2023 when participants were aged 25 to 28 years. Participants with available genetic data and at least one quantitative symptom score included.

**Main outcomes and measures:** Quantitative symptom scores and self-reported psychiatric diagnoses. Associations were tested using generalized estimating equations for three types of polygenic scores: uncorrected polygenic scores; a transdiagnostic polygenic score indexing shared genetic variance across 11 psychiatric conditions (*p*); and residual disorder-specific scores corrected for *p* (non-*p*).

**Results:** Analyses included between 5,789 and 6,546 (mean [SD] age, 26.4 [0.93] years; 7,244 [51.6%] female). The polygenic score for *p* consistently showed stronger associations with symptom scores than uncorrected polygenic scores, while associations with self-reported diagnoses were similar. Most uncorrected polygenic scores exhibited extensive cross-trait associations, which were substantially attenuated accounting for *p*, suggesting that much of the genetic signal captured by polygenic scores reflects transdiagnostic liability. Some non-p polygenic scores retained associations with their corresponding-traits, indicating residual specificity.

**Conclusions and relevance:** In this population-based sample of young adults, associations between polygenic scores and psychopathology outcomes primarily reflect transdiagnostic genetic risk, with limited evidence of disorder-specificity. Accounting for transdiagnostic genetic liability could improve the specificity and interpretability of polygenic scores.

**Key Points:** Question: To what extent do polygenic scores for psychiatric disorders reflect transdiagnostic (p) versus disorder-specific genetic risk across psychopathology?

Findings: In a UK population-based cohort of approximately 6,000 individuals, polygenic scores for psychiatric disorders were associated with both quantitative symptom scores and self-reported diagnoses largely due to the transdiagnostic *p*, with limited evidence of disorder-specificity.

Meaning: Accounting for transdiagnostic genetic risk could improve the specificity and interpretability of polygenic scores in psychiatric research and practice.

## Introduction

Genome-wide association studies (GWASs) have transformed our understanding of the genetic architecture of psychiatric disorders.^1^ In addition to identifying replicable genetic loci associated with disorders, summary statistics from GWASs made it possible to construct polygenic scores (PGSs), which aggregate the effects of thousands of single nucleotide polymorphisms (SNPs) identified in a GWAS to estimate an individual’s genetic liability for a given trait or disorder.^2^

Despite the increasing use of PGSs in psychiatric research,^3^ their clinical utility remains limited.^4^ One major constraint is their modest explanatory power, as PGSs capture only a fraction of SNP heritability. This limitation stems partly from underpowered GWAS sample sizes, unmeasured genetic variants (e.g., rare variants), phenotypic heterogeneity, and imprecise trait definitions. Another major limitation is the lack of specificity in PGSs for the disorders for which they were derived. PGSs show transdiagnostic associations with multiple psychiatric outcomes, reflecting substantial genetic overlap across disorders.^5–7^ Although pairwise genetic correlations across disorders are substantial, they remain well below 1.0, suggesting both shared and unique genetic contributions. However, the extent to which PGSs capture disorder-specific versus transdiagnostic genetic liability remains unclear.

One approach to investigating genetic specificity is to create PGSs corrected for transdiagnostic effects. Shared variance across psychiatric disorders is often conceptualized as a general factor of psychopathology, or *p*, which represents broad liability to multiple mental health conditions.^8–10^ Although initially identified at the phenotypic level, the *p* factor has since been observed at genetic and genomic analyses ^11–14^.

In prior work,^13^ we used genomic structural equation modelling (genomic SEM)^15^ to decompose the genetic variance across 11 major psychiatric disorders into a transdiagnostic (*p*) and disorder-specific residual components. This yielded summary statistics for both the shared and unique genetic components (referred to as *non-p* to represent residual genetic variance independent of shared liability).

In the present study, we used these summary statistics to construct PGSs in a UK population-based cohort of young adults from the Twins Early Development Study. We tested associations between quantitative symptom scores and self-reported diagnoses and three types of PGSs: uncorrected PGSs, a transdiagnostic PGS indexing shared genetic liability across 11 disorders (p), and disorder-specific PGSs that are corrected for shared variance (non-p). By comparing these PGSs, we aimed to evaluate the extent to which PGSs reflect transdiagnostic or disorder-specific genetic risk. This study was preregistered with the Open Science Framework prior to accessing the data (OSF; [LINK](https://osf.io/29xyt)).

## Methods

### Sample

Participants were drawn from the Twins Early Development Study (TEDS)^16^, a longitudinal cohort of 13,759 twin pairs (27,518 individuals) born in England and Wales between 1994-1996. The original cohort was representative of the UK population in terms of ethnicity and socio-economic status^17^, which was largely maintained at age 26, despite some attrition^16^.

Mental health data were collected between 2021 and 2023, when participants were aged 25- to 28 years, via self-report questionnaires administered online or by post.

Genotyping was conducted on two different platforms (AffymetrixGeneChip 6.0 and Illumina HumanOmniExpressExome-8v1.2). Details on DNA collection, genotyping, and quality control are described elsewhere^14,18^. Individuals with both genetic data and at least one quantitative symptom score were included in the present study. Final analytic samples ranged from 5,789 to 6,546 individuals (including DZ twins), depending on the outcome measure. TEDS has ethical approval from King’s College London Research Ethics Committee (PNM/09/10-104) and written informed consent was obtained from all participants.

### Measures

#### Quantitative Symptom Measures

Symptom data were available for eight psychiatric disorders (Table 1). Descriptions of each measure are provided in eMethods (Supplement 1), with the descriptive data in eTable 1 (Supplement 2).

**Table 1.** Quantitative symptom measures used in the current study. . ANX = Anxiety disorder; ADHD = Attention-deficit/hyperactivity disorder; ASD = Autism spectrum disorder; MDD = Major depressive disorder; BIP = Bipolar disorder; ALCH = Problematic alcohol use; PTSD = Post-traumatic stress disorder; SCZ = Schizophrenia. Symptoms related to SCZ were assessed using dimensional measures of psychotic-like experiences, including paranoia and hallucinations.

| Symptoms (related disorder) | Measure | Reference |
| --- | --- | --- |
| Anxiety symptoms (ANX) | Generalized Anxiety Disorder- Dimensional (GAD-D) | 19 |
| Attention-deficit symptoms (ADHD) | Conners Rating Scale | 20 |
| Autistic traits (ASD) | Ritvo Autism and Asberger Diagnostic Scale (RAADS-14) | 21 |
| Depressive symptoms (MDD) | Moods and Feelings Questionnaire (MFQ-13) | 22 |
| Mania/Hypomania symptoms (BIP) | Mood Disorder Questionnaire (MDQ) | 23 |
| Alcohol use (ALCH) | Alcohol Use Disorder Identification Test (AUDIT) | 24 |
| Post-traumatic stress disorder symptoms (PTSD) | Post-traumatic Stress Disorder Checklist (PCL-6) | 25 |
| Schizophrenia (SCZ) | Specific Psychotic Experiences Questionnaire (SPEQ): | 26 |
|  | Hallucinations and Paranoia subscales |  |

### Self-Reported Diagnoses

Self-reported diagnostic data were available for nine disorders: attention-deficit/hyperactivity disorder (ADHD), anorexia nervosa (AN), anxiety disorder (ANX), autism-spectrum disorder (ASD), bipolar disorder (BIP), major depressive disorder (MDD), obsessive-compulsive disorder (OCD), psychosis/psychotic illnesses (excluding schizophrenia; SCZ), and post-traumatic stress disorder (PTSD). Participants were provided with a list of mental health conditions and asked whether they had ever received a professional diagnosis, regardless of their current status at the time of data collection. Responses were recorded as yes/no.

Frequencies are presented in eTable 2 (Supplement 2). Due to lack of SCZ cases, psychosis/psychotic illnesses were used as a proxy in analyses involving the SCZ PGS.

### GWAS summary statistics

We used publicly available summary statistics for 11 major psychiatric disorders: ADHD ^27^, SCZ ^28^, ANX ^29^, ASD ^30^, BIP ^31^, MDD ^32^, PTSD ^33^, Tourette’s syndrome (TS) ^34^, problematic alcohol use (ALCH) ^35^, OCD ^36^, and AN ^37^. All GWASs were based on case-control designs, except for ALCH, which used symptom-level measure of alcohol use. Details are provided in eTable 3 (Supplement 2). Using genomic SEM, we derived summary statistics for a transdiagnostic factor (p) and residual disorder-specific components (hereafter *non-p*) ^13^. These summary statistics were used to construct PGSs in the current study.

### Polygenic score construction in TEDS

We constructed PGSs in the TEDS sample using LDPred2^38^ with the ‘auto’ option and default parameters. Analyses were restricted to HapMap3 variants^39^ and based precomputed linkage disequilibrium (LD) matrices from the UK Biobank^39^. Both the discovery GWASs and the target sample were of European ancestry.

### Statistical Analysis

Analyses were conducted using R (v.4.4.2)^40^ and reported following the Strengthening the Reporting of Observational Studies in Epidemiology (STROBE) reporting guideline (eTable 4, Supplement 2). Figure 1 outlines our analytic framework.

**Figure 1.**
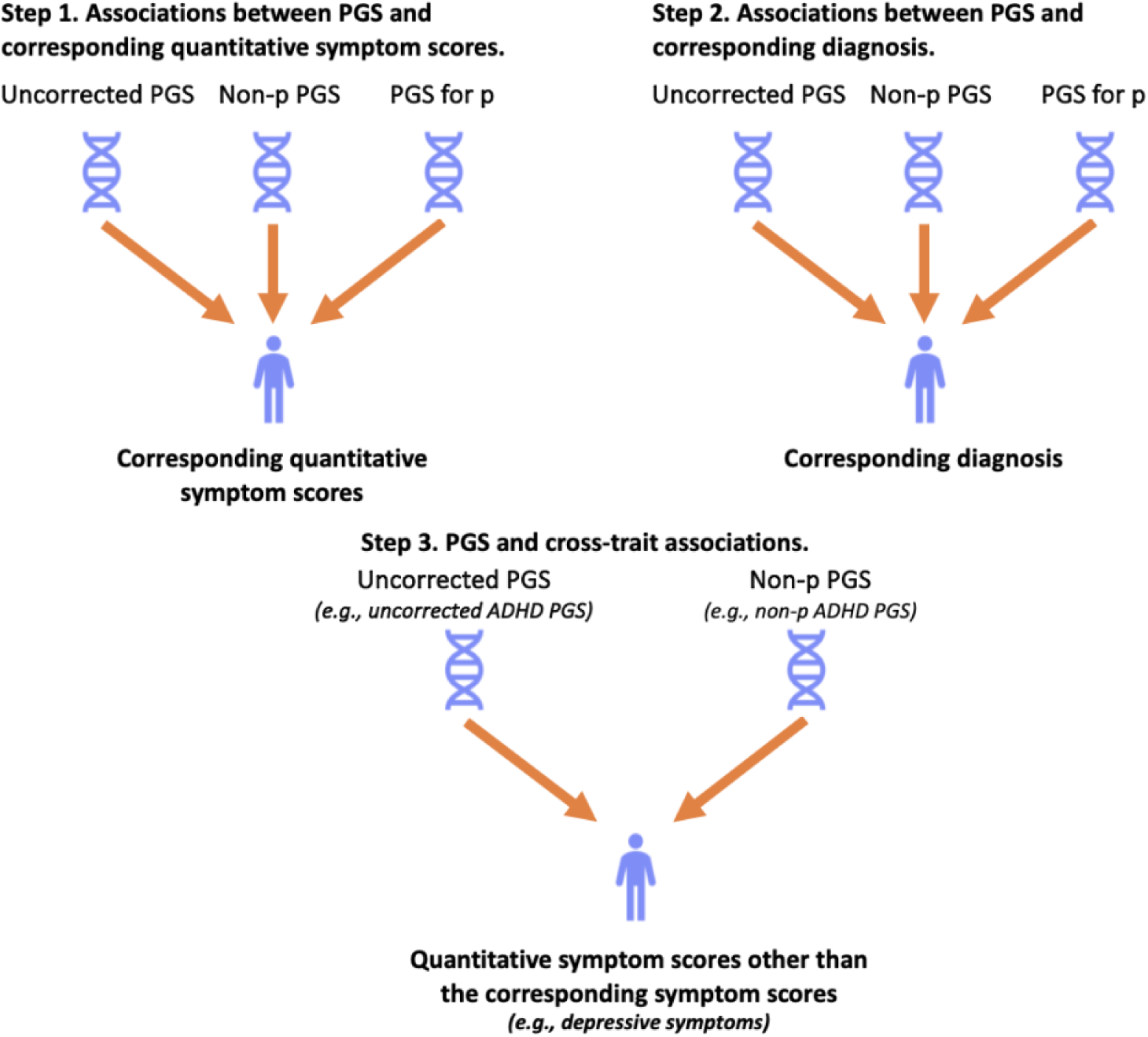
Three analytic steps. The figure illustrates the three-step analysis. In each step, PGSs were tested independently, one at a time, in separate models. For example, in Step 1, the uncorrected PGS was tested first, followed by separate tests for the non-p PGS and the PGS for p. In Step 1, we assessed associations between each PGS and its corresponding quantitative symptom score. In Step 2, we examined associations between each PGS and its corresponding diagnosis. Finally, in Step 3, we investigated cross-trait associations, such as uncorrected and non-p ADHD PGSs predicting depressive symptom scores.

We first examined associations between each PGS and its corresponding symptom score and self-reported diagnosis. To assess pleiotropy, we tested cross-trait associations between each PGS and symptom scores beyond its primary phenotype. Analyses were repeated after excluding individuals with self-reported diagnoses to assess whether associations were primarily driven by these individuals.

To test robustness, we conducted sensitivity analyses comparing symptom severity and diagnostic burden in individuals in the top 1% of the p PGS distribution relative to the uncorrected and non-p PGSs. We also examined associations between the p PGS and high-risk individuals (based on clinical cutoffs), as well as between the top versus bottom deciles of symptom distributions (eMethods in Supplement 1).

All regressions were performed using generalized estimating equations (GEE), with an exchangeable correlation structure, to account for relatedness within the sample. Continuous outcomes were residualized for age and sex and standardized prior to analysis. All models included the top 10 ancestry PCs, genotyping batch, and chip as covariates. Gaussian models were applied for continuous outcomes, and binomial models for binary outcomes, using R package *gee* ^41^. To account for multiple testing, false discovery rate (FDR) corrections were applied (FDR 5%) ^42^.

## RESULTS

### Associations between PGSs and corresponding quantitative symptom scores

Figure 2 presents the associations between PGSs and corresponding symptom scores. Four key findings emerged: 1) All uncorrected PGSs were significantly associated with their corresponding symptom scores, although effect sizes were modest. 2) Non-p PGSs generally showed attenuated associations compared to uncorrected PGSs. Exceptions were observed for alcohol use and PTSD, which retained significant associations, suggesting some disorder-specific genetic signal. The p PGS was significantly associated with all symptom scores except alcohol use, with the strongest association with PTSD symptoms. In five of nine comparisons, the p PGS outperformed the uncorrected PGS, and performed comparably in the remaining four. 4) These patterns were consistent after excluding individuals with self-reported diagnoses, although some associations were attenuated and no longer statistically significant. Full results are presented in eTables 5 and 6 in Supplement 2.

**Figure 2.**
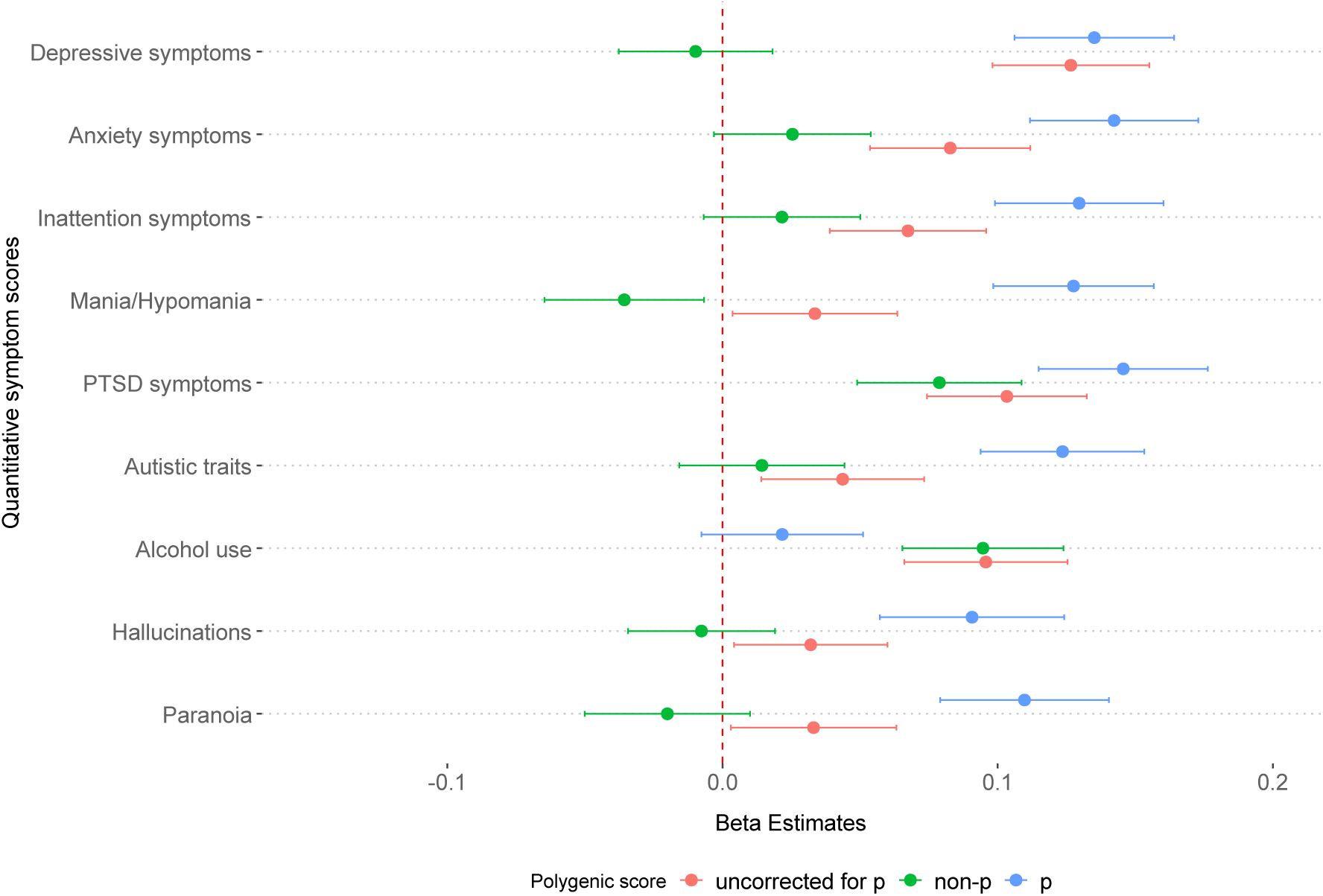
Associations between PGSs (uncorrected, transdiagnostic (p) and non-p), and corresponding quantitative symptom scores. (e.g., MDD PGS predicting depressive symptoms) for uncorrected PGSs (blue), PGSs corrected for p (non-p; green), and PGS for p (purple). Bars represent the effect size of standardized regression coefficients, error bars indicating 95% confidence intervals. All variables were residualized for age and sex, and the residuals were standardized before regression analyses. All analyses were adjusted for the first 10 genetic principal components and genotyping chip and batch. Exact estimates and *P* values are reported in <u>eTable 5</u>.

### Associations between PGSs and corresponding self-reported diagnoses

Figure 3 presents odds ratios for associations between each PGS and its corresponding self-reported diagnosis (details in eTable 7 in Supplement 2). Despite the relatively small number of individuals with self-reported diagnoses, seven of nine uncorrected PGSs were significantly associated with their respective diagnoses. Notably, non-p PGSs for AN and PSTD also retained significant associations, suggesting the presence residual disorder-specific genetic variance. The p PGS showed comparable associations to the uncorrected PGSs but did not outperform them.

**Figure 3.**
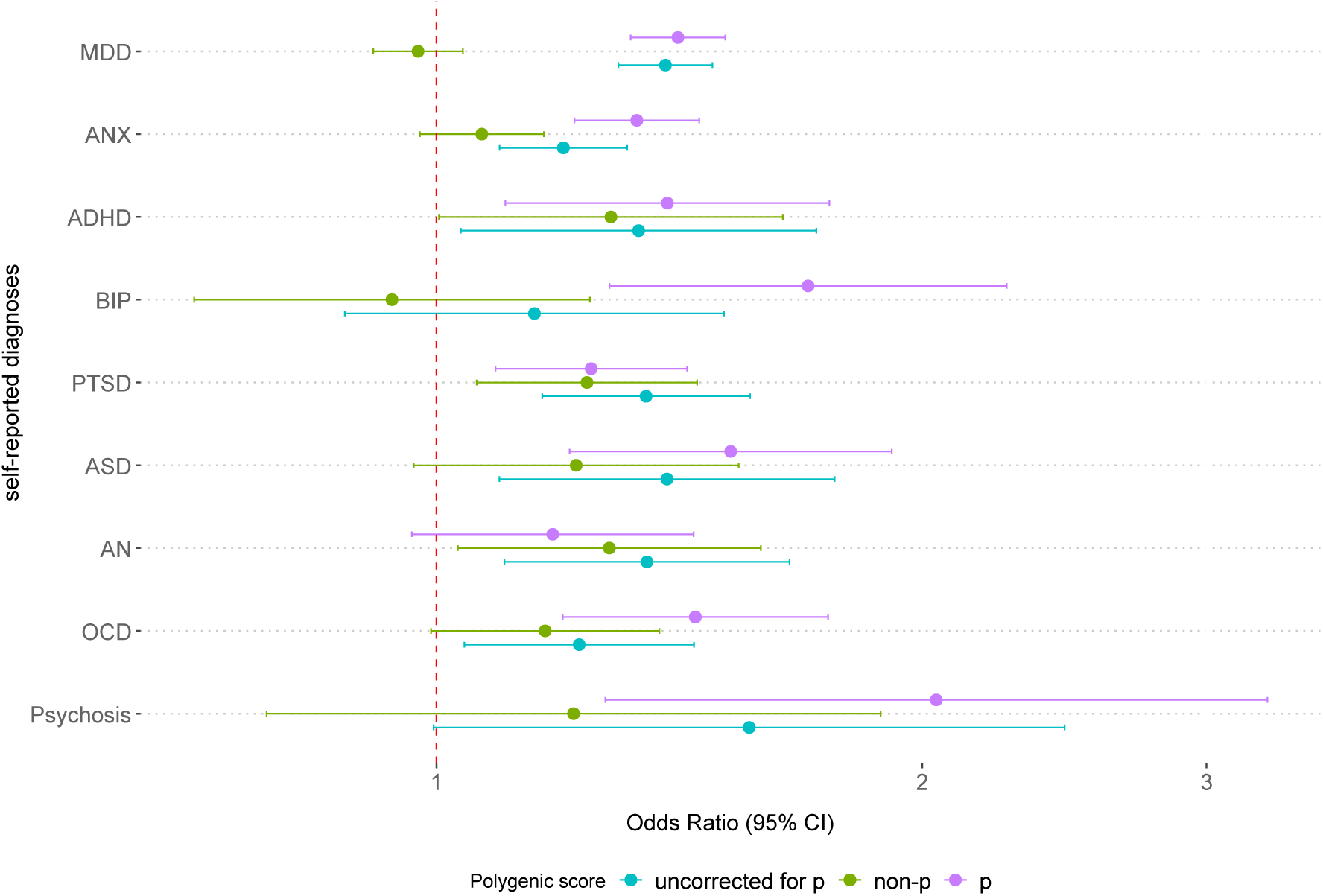
Associations between PGSs (uncorrected, transdiagnostic (p) and non-p), and corresponding self-reported diagnoses. (e.g., MDD PGS predicting MDD diagnosis), for uncorrected PGSs (blue), non-p PGSs (green), and the PGS for p (purple), represented as odds ratios (OR) with 95% confidence intervals (CI). The dashed line indicates the null value (OR = 1). Self-reported diagnoses were available for nine disorders: MDD = major depressive disorder; ANX= anxiety disorder; ADHD = attention-deficit/hyperactivity disorder; BD = bipolar disorder; PTSD = post-traumatic stress disorder; ASD= autism-spectrum disorder; AN = anorexia nervosa; OCD = obsessive-compulsive disorder. PGS for SCZ was used to predict the diagnosis of psychosis/ psychotic illnesses. All analyses were adjusted for the first 10 genetic principal components, genotyping chip and batch. Exact estimates and *P* values are reported in eTable 7.

### Cross-trait associations between PGSs and quantitative symptom scores

Cross-trait analyses (eTable 8 in Supplement 2) revealed extensive pleiotropy. Several uncorrected PGSs, including those for ANX, MDD, SCZ, PTSD, and ADHD showed broad cross-trait associations across multiple non-corresponding symptom scores. In many cases, cross-trait associations were as strong as, or stronger than, same-trait associations. For example, the MDD PGS was similarly associated with symptoms of MDD (*β* = 0.13, SE = 0.01), ASD (*β* = 0.13, SE = 0.01), and PTSD (*β* = 0.14, SE = 0.01). An exception was the ALCH PGS, which was associated only with symptoms of ADHD and MDD. Accounting for transdiagnostic variance substantially reduced most associations, indicating that much of the cross-trait signal reflected shared genetic risk. Results were similar, but attenuated, after excluding individuals with self-reported diagnoses (eTable 9 in Supplement 2).

### Sensitivity analyses

Sensitivity analyses supported the robustness of the primary findings. First, individuals in the top 1% of the p PGS distribution showed elevated symptom levels across multiple domains and higher diagnostic comorbidity compared to individuals in the top 1% of uncorrected and non-p PGS distributions (eFigure 1, Supplement 1), consistent with a broad phenotypic expression of transdiagnostic liability. Second, to assess clinical relevance, we tested the association between each PGS and high-risk status and compared individuals in the top and bottom deciles of the symptom distributions. The p and uncorrected PGSs performed similarly in identifying individuals with elevated symptoms, both showing stronger associations with high-risk status (eFigure 2, Supplement 1) and greater differentiation between the extremes of the distributions (eFigure 3, Supplement 1). Non-p PGSs generally showed weaker associations, with the exception of ALCH and PTSD. Full results are provided in eResults (Supplement 1) and eTables 10-12 (Supplement 2).

## DISCUSSION

In this population-based sample of young adults, we examined whether polygenic scores (PGSs) for major psychiatric disorders primarily index transdiagnostic or disorder-specific genetic liability. Using both quantitative symptom scores and self-reported diagnoses, we compared three types of PGSs: uncorrected PGSs, a transdiagnostic PGS (p), and disorder-specific PGSs corrected for transdiagnostic effects (non-p). Overall, associations between PGSs and psychopathology outcomes were primarily driven by transdiagnostic genetic risk, with limited evidence of disorder-specificity. To our knowledge, this is the first study to systematically compare the different types of PGSs across a range of psychiatric outcomes using both quantitative and (self-reported) diagnostic data.

These findings highlight the importance of accounting for transdiagnostic liability when interpreting associations between PGSs and psychopathology. The p PGS was consistently associated with symptom scores, often outperforming uncorrected PGSs for corresponding domains. Cross-trait analyses revealed widespread pleiotropy, and adjusting for p substantially attenuated associations between PGSs and non-corresponding symptoms, suggesting that may uncorrected PGS associations reflect transdiagnostic genetic risk. These results align with accumulating evidence from genomic studies demonstrating extensive overlap in the genetic architecture of psychopathology.^5,7,12,13^ Without accounting for this shared variance, associations between PGSs and psychiatric outcomes may be misinterpreted as disorder-specific effects.

Although the p PGS likely benefits from increased statistical power due to the aggregation of GWAS data across 11 disorders, its pattern of associations suggests that power alone does not account for its performance. Instead, the extent to which a PGS reflects transdiagnostic versus disorder-specific liability partly depends on the phenotypic granularity of the discovery GWAS.^43^ GWASs based on narrowly defined or dimensional phenotypes tend to yield greater specificity.^43,44^ This was reflected in our findings for problematic alcohol use: the ALCH PGS, derived from an item-level GWAS ^35^ and assessed with the same measure in our sample, was the only PGS – both uncorrected and non-p – that was almost exclusively associated with its corresponding phenotype. This likely reflects both the phenotypic precision of the discovery GWAS and the observable nature of behaviour related to problematic alcohol use (e.g., failing to complete a task due to drinking).

Moreover, the p PGS showed stronger associations than uncorrected PGSs with quantitative symptom measures, but not with self-reported diagnoses. If statistical power was the main driver of this effect, similar advantages would be expected across both types of outcomes.

Instead, the comparable performance of the p and uncorrected PGSs for diagnostic outcomes suggests that uncorrected PGSs retain some disorder-specific signal, potentially due to their derivation from case-control GWASs that align more closely with diagnostic phenotypes. In contrast, the stronger associations between the p PGS and symptom scores likely reflect better alignment with dimensional, transdiagnostic measures that capture heterogeneity within and overlapping symptoms across disorders. Supporting this, when symptom scores were dichotomized to approximate clinical thresholds, the performance of the p and uncorrected PGSs converged, suggesting that uncorrected PGSs may better capture disorder-specific risk under more clinically relevant cases.

Despite the dominance of transdiagnostic genetic risk, some non-p PGSs retained significant associations with their corresponding symptom scores and self-reported diagnoses, despite the limited number of cases. For example, the non-p PGS for PTSD was modestly but robustly associated with both PTSD symptoms and diagnoses, despite only 38% of its genetic variance being independent of the p factor.^13^ This suggests that, while transdiagnostic effect account for much of the observed genetic signal, unique liability remains detectable, particularly for biologically cohesive phenotypes such as PTSD.^45^ As GWAS sample sizes grow and phenotyping becomes more refined, these residual disorder-specific signals may become more informative. Emerging methods offer promising avenues for improving the specificity of PGSs.^46,47^ In the meantime, p-corrected summary statistics, like those used in our study, offer a practical alternative for investigating specificity.

Extending prior research,^32,35,48,49^ our findings demonstrate shared genetic risk between clinical diagnoses and corresponding symptoms in the general population. This is aligned with dimensional frameworks such as RDoC ^50^ and HiTOP ^51^, which conceptualize psychopathology along a continuum of severity rather than as discrete categories. These frameworks more accurately reflect the shared symptomatology and highly polygenic nature of psychiatric traits. Quantitative symptom measures, which capture variation across the full spectrum of severity, may therefore be particularly well-suited to capturing transdiagnostic risk. When paired with a transdiagnostic PGS such as the p PGS, these measures may offer a complementary approach for population-level risk stratification^51–53^ and informing transdiagnostic interventions.^54^

## Limitations

Several limitations should be noted. First, reliance on self-report data may introduce recall or reporting bias.^55^ The use of composite symptom scores may also oversimplify the complexity of psychopathology. Alternative methods, such as network analysis, could better capture the complexity of mental health problems.^56,57^ Second, analyses were restricted to individuals of European ancestry and the target sample consisted of younger adults, limiting the generalizability of our findings to other ancestral populations and age groups. Third, while our sample size is modest for detecting associations with less prevalent disorders, the TEDS cohort offers rich phenotypic data. Future research would benefit from replicating these findings in larger and more diverse populations with similarly detailed measures.

An additional consideration is that the PGSs for p and non-p were based on genomic SEM-derived summary statistics.^13^ While SEM is a powerful framework for modelling complex data, the resulting factors are not causal entities ^58^ and are sensitive to constructs included in the model.^59^ Consequently, the p factor, and by extension non-p, reflects the structure and limitations of the underlying GWASs, including phenotype definitions and ascertainment strategies ^44,60^. For example, the inclusion of individuals with comorbid diagnoses in GWASs may inflate genetic correlations,^61^ potentially amplifying p. However, simulations showed that extremely high rates of diagnostic overlap and misclassification are required to account for the observed genetic overlaps.^5^ Additionally, non-p PGSs were constructed by statistically removing p-related variance, but this process is imperfect. Therefore, the p factor should not be viewed as a pure index of general psychopathology, but rather as a data-driven summary of genetic covariance across GWASs. Similarly, non-p PGSs approximate residual trait-specific liability rather than representing pure indicators of specificity. Together, these PGSs offer a complementary approach to current GWAS methods for investigating specificity and transdiagnostic genetic liability.

## Conclusion

Using both quantitative symptom measures and self-reported diagnoses, our findings indicate that polygenic scores derived from current GWASs of psychiatric disorders primarily reflect transdiagnostic genetic liability, with limited evidence of disorder-specificity. Accounting for the transdiagnostic liability may improve the specificity and interpretability of PGSs for specific disorders, while transdiagnostic PGS may provide opportunities for risk stratification and treatment.

## Data Availability

The TEDS resource is held by King's College London. Data can be made available, subject to a data sharing agreement as detailed at https://www.teds.ac.uk/researchers/teds-data-access-policy.

