## Supplementary material for "Specificity of polygenic scores for psychiatric disorders beyond transdiagnostic genetic risk (p)": Online Supplement 1

#### Supplemental Online Content

**eMethods**
**eResults**
**eFigure 1. Heatmap of standardized symptom scores in the top 1% of each polygenic score distribution.**
**eFigure 2. Odds ratios (ORs) for high-risk group status by polygenic scores.**
**eFigure 3. Odds ratios (ORs) for top versus bottom symptom deciles by polygenic score.**
**eReferences**

### eMethods

**Psychopathology measures**
Questionnaires used to measure symptoms associated with psychiatric disorders. The questionnaires are described in further detail in the TEDS data dictionary (https://www.teds.ac.uk/datadictionary/home.htm).

Anxiety symptoms:
Anxiety symptoms were measured using the Generalised Anxiety Disorder – Dimensional (GAD-D) [1]. This scale consists of 10 items that assess thoughts, feelings, and behaviours, commonly associated with concerns about family, health, finances, school, and work. Each item is rated on a 5-point scale: "Never" (0), "Occasionally" (1), "Half of the time" (2), "Most of the time" (3), "All of the time" (4). The total score ranges from 0 to 40, with higher scores indicating greater levels of anxiety.

Attention Deficit Hyperactivity Disorder (ADHD) symptoms:
ADHD symptoms were measured using the Conners-3-Self-Report [2], which assesses ADHD symptoms in adults. For this study, only the 11 items related to inattention were included. These items measure difficulties such as poor concentration, trouble maintaining focus, being easily distracted, and difficulty starting and/or finishing tasks. Responses are rated on a 4-point scale: "Not true at all" (0), "Somewhat true" (1), "Mainly true" (2), and "Definitely true" (3). The total score ranges from 0 to 33, with higher scores indicating greater levels of attention deficits.

Alcohol use:
Alcohol use was assessed using the adapted version of the Alcohol Use Disorders Identification Test (AUDIT) [3], a 10-item screening tool used to assess alcohol consumption, behaviours, and problems related to alcohol use. The first three items of the scale measure general consumption (i.e., quantity and frequency of drinking) as well as one item measuring frequency of heavy/binge drinking. The items 4-10 focus on the problematic consequences of drinking. The responses are rated on a 5-point scale, indicating the severity or frequency of the behaviors. The total score is the sum of the values from each of the 10 items and ranges from 0 to 40, with higher scores indicating more serious alcohol-related problems.

Autism Spectrum Disorder (ASD) traits:
Autistic traits were assessed using the Ritvo Autism and Asperger Diagnostic Scale (RAADS-14) [4], a screening tool for ASD in adults. The measure evaluates persistent ASD symptoms across three subdomains: mentalizing deficits, social anxiety, and sensory reactivity. In this study, the questionnaire was reduced from 14 to 6 items, retaining 3 socio-communicative and 3 non-social items. Each item is rated as: "True now and when I was young" (0), "True only now" (1), "True only when I was younger than 17" (2), and "Never true" (3). After reverse-coding the items, the total score ranges from 0 to 18, with higher scores indicating more pronounced autistic traits.

Depression symptoms:
Depression symptoms were assessed using the Short Mood and Feelings Questionnaire (MFQ-13) [5]. This 13-item scale aims to capture the presence of depressive symptoms over the last two weeks. Each item is rated as "Not true" (0), "Sometimes true" (1), and "True" (2). The total score ranges from 0 to 26, with higher scores indicating more severe depressive symptoms.

Mania/Hypomania symptoms:
Symptoms of mania and hypomania were assessed using the Mood Disorder Questionnaire (MDQ) [6], a self-report screening tool for bipolar disorder. The MDQ includes questions about presence of bipolar symptoms, symptom co-occurrence and impaired functioning. It consists of 13 yes/no items which relate to the presence of symptoms based on DSM-IV [7] criteria for a hypomanic or manic episode. Additional items inquire whether symptoms occur during the same period and affect functioning. The total score is the sum of the symptoms, ranging from 0 to13.

Post-Traumatic Stress Disorder (PTSD) symptoms:
Current PTSD symptoms were assessed using the Post-Traumatic Stress Disorder Checklist (PCL-6) [8], a validated short form of the 17-item PCL [9]. The PCL-6 consists of 6 items that measure the clusters of PTSD as described in DSM-V [10]: re-experiencing, avoidance, negative cognitions and hyperarousal- and a total symptom severity score (PCL-Total) as the sum of those sub-phenotypes. In the TEDS sample, participants were asked to respond to the PCL-6 items in reference to *any* stressful experience. Items are rated on a 5-point scale: "Not at

all" (0), "A little bit" (1), "Moderately" (2), "Quite a bit" (3), "Extremely" (4), with the total score ranging from 0 to 24.

Psychotic symptoms:
Two key types of experiences associated with psychosis – paranoia and hallucinations – were measured using the subscales of Specific Psychotic Experiences Questionnaire (SPEQ) [11]. The paranoia subscale consists of 15 items, and the hallucinations subscale consists of 9 items. Responses are rated on a 7-point scale: "Not at all" (0), "Rarely" (1), "Once a month" (2), "Once a week" (3), "Several times a week" (4), and "Daily" (5). The total score for the paranoia subscale ranges from 0 to 75, and the total score for the hallucinations subscale ranges from 0 to 45.

**Sensitivity analyses**
We conducted a series of sensitivity analyses to ensure robustness of the primary findings. First, to assess what the p PGS capture phenotypically in the current sample, we examined whether individuals in the top 1% of the p PGS distribution showed distinct patterns of symptom severity and diagnostic burden compared with those in the top 1% of each uncorrected and non-p PGS. We also examined self-reported diagnoses and comorbidity patterns.

Second, we assessed the clinical relevance of PGSs, we compared associations across the p PGS, uncorrected PGSs, and non-p PGSs using two complementary approaches. First, we defined binary high-risk groups for each of the symptom score distribution using either established clinical cutoffs (for MDD [12,13], ANX [14], PTSD [15], ALCH, and hypomania) or the top 5% of the distribution (for ADHD, ASD) when clinical cutoffs were not available. The remaining sample served as the control group. Next, to evaluate performance across the full spectrum of severity, we also compared individuals in the top versus bottom 10% of each quantitative symptom score distribution.

For both analyses, logistic regressions were conducted using a generalized estimating equations (GEE) framework. All continuous variables were residualized for age and sex, and the residuals were standardized prior to analyses. All models included the same covariates as in the primary analyses (i.e., top 10 ancestry PCs, genotyping batch, and chip). Effect sizes are presented as adjusted odds ratios (ORs) per standard deviation (SD), with 95% confidence intervals.

#### eResults

##### Sensitivity analyses

**Phenotypic profile of individuals in the top 1% of the p PGS**
Individuals in the top 1% of the p PGS exhibited elevated symptom levels across nearly all psychiatric domains, consistent with the interpretation of the p as an index of transdiagnostic liability (eFigure 1). In contrast, those in the top 1% of uncorrected PGSs showed more heterogeneous elevations across symptom profiles, with generally lower symptom elevations. Notable exceptions included the PGSs for ADHD and ALCH PGS, which showed broader symptom elevations relative to other uncorrected scores. Most non-p PGSs yielded average symptom levels close to the sample mean, indicating more limited phenotypic impact.

Rates of self-reported diagnoses were similar across the top 1% of all PGSs. For example, in the top 1% of the p PGS distribution 47 of 51 individuals (92.2%) reported at least one psychiatric diagnosis. The most frequently endorsed conditions were MDD (23 cases; 47.1%), followed by OCD (7 cases; 13.7%), ANX (6 cases; 11.8%), and PTSD (4 cases;7.8%). Results were similar for uncorrected PGSs, including ADHD (49 cases), MDD (40 cases), and ANX (36 cases), and for non-p PGSs, such as ADHD non-p (35 cases). Results are provided in eTable 10 in Supplement 2. A slightly different pattern emerged when examining individuals reported two or more diagnoses: the p (11 individuals), ADHD (13 individuals), and ANX (10 individuals).

It is important to note that the top 1% PGS groups are not independent. For example, an individual with high p PGS and 2+ diagnoses likely also fall into the top 1% for ADHD or ANX. Therefore, these results illustrates the general patterns of psychiatric burden among individuals with high PGSs.

**PGS associations with high-risk vs controls**
The p PGS was significantly associated with high-risk group across nearly all domains tested (ORs ranging from 1.26 to 1.45; all $p < .001$), with the exception of alcohol use symptoms (OR = 1.11, 95% CI [0.88–1.41]). Uncorrected PGSs were also significantly associated with high-risk status in most domains, with ORs very similar to p PGSs. In most domains, non-p PGSs were not associated with high-risk status. Consistent with primary findings, exceptions included the PTSD non-p PGS (OR = 1.18, 95% CI [1.07–1.30]) and ALCH non-p PGS (OR = 1.44, 95% CI [1.16–1.79]), suggesting retained disorder-specificity. Results are provided in eTable 11 in Supplement 2.

**PGS associations with top 10% vs bottom 10% of symptom score distributions**
Consistent patterns of associations were observed when comparing individuals in the top versus bottom deciles of symptom severity distributions. The p PGS and uncorrected PGSs showed significant associations across most domains. In addition to non-p PGS for PTSD and ALCH, non-p ADHD also showed a modest but significant association with extremes (OR = 1.18, 95% CI, 1.03–1.34) were significantly associated with symptom extremes. Results are provided in eTable 12 in Supplement 2.

### eFigures

**eFigure 1. Heatmap of standardized symptom scores in the top 1% of each polygenic score distribution.** Rows represent types of PGS, columns represent symptom domains. Colour intensity reflects the mean standardized symptom score within each top 1% polygenic score group, with red indicating above-average symptom levels and blue indicating below-average levels. BIP: Bipolar disorder; MDD: Major depressive disorder; SCZ: Schizophrenia; ASD: Autism-spectrum disorder; ANX: Anxiety disorder; PTSD: Post-traumatic stress disorder; ALCH: Problematic alcohol use.

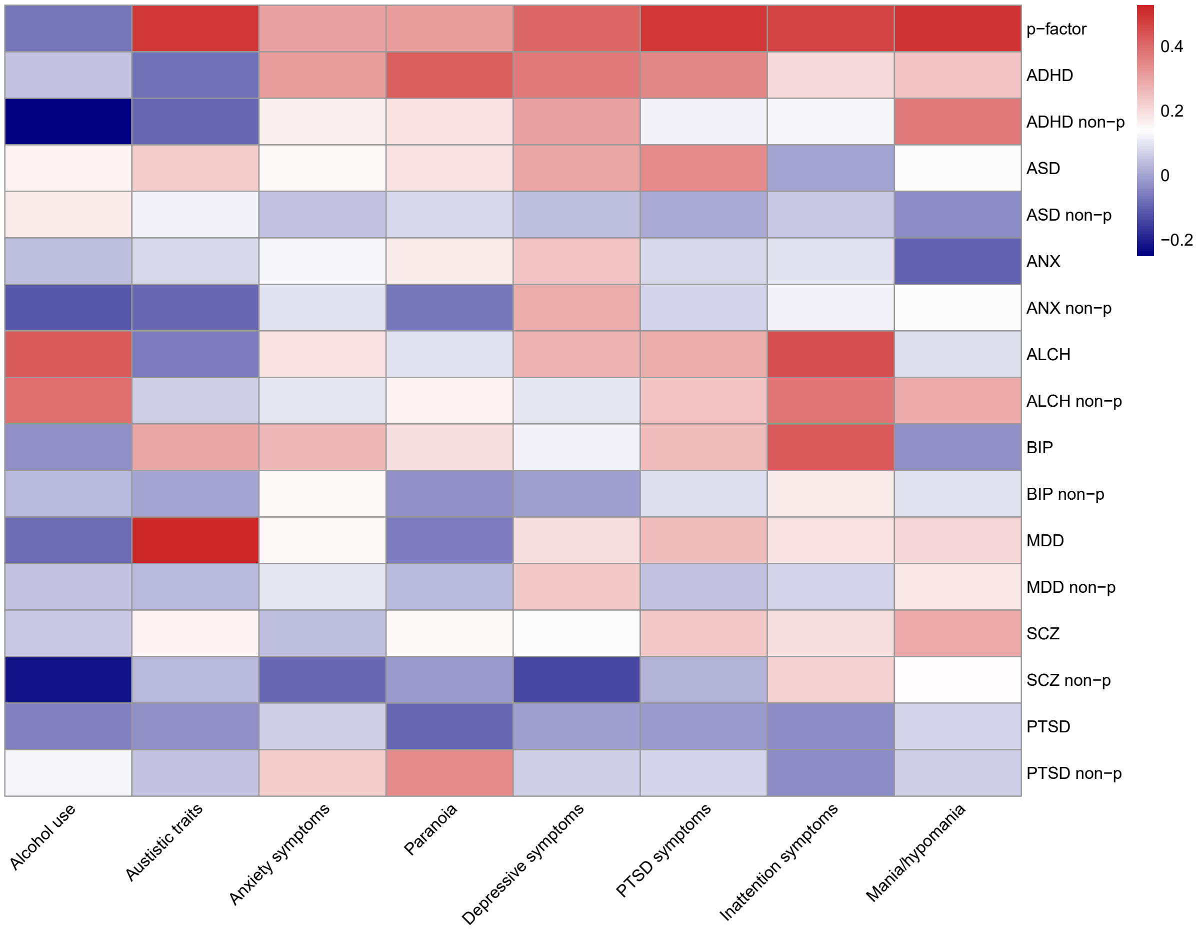

**eFigure 2. Odds ratios (ORs) for high-risk group status by polygenic scores.** Each point represents the odds ratio (OR) for being in the high-risk group, as estimated from logistic regression models within generalized estimating equations. Models were run separately for each of three polygenic scores: transdiagnostic (p), uncorrected, and p-corrected (non-p). All models included the first 10 principal components, genotyping batch, and chip as covariates. Error bars indicate 95% confidence intervals. The dashed line indicates the null value (OR = 1). The x-axis is plotted on a log scale.

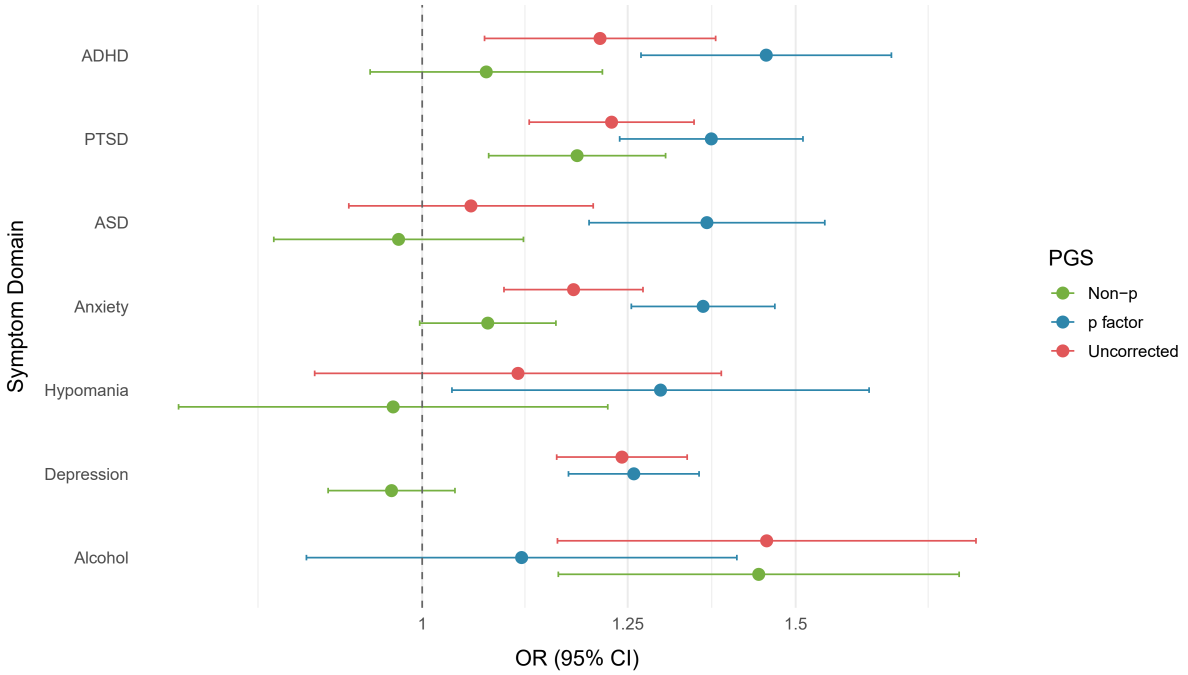

**eFigure 3. Odds ratios (ORs) for top versus bottom symptom deciles by polygenic score.** Each point represents the odds ratio (OR) for being in the top 10% versus bottom 10% of symptom severity, as estimated from logistic regression models within generalized estimating equations. Models were run separately for each of three polygenic scores: transdiagnostic (p), uncorrected, and p-corrected (non-p). All models included the first 10 principal components, genotyping batch, and chip as covariates. Error bars indicate 95% confidence intervals. The dashed line indicates the null value (OR = 1). The x-axis is plotted on a log scale.

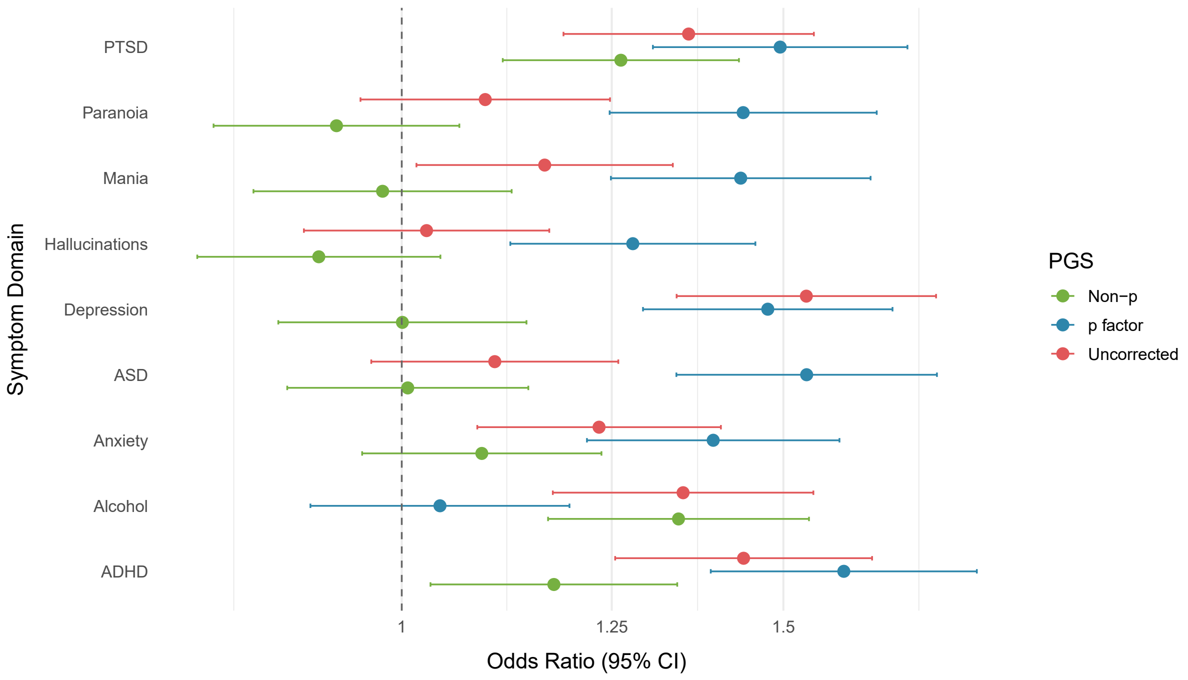
